# Deep learning application to automatic classification of pharmacist interventions

**DOI:** 10.1101/2022.11.30.22282942

**Authors:** Ahmad Alkanj, Julien Godet, Erin Johns, Bénédicte Gourieux, Bruno Michel

## Abstract

**Background:** Pharmacist Interventions (PIs) are actions proposed by pharmacists during the prescription review process to address non-optimal drug use. PIs must be triggered by drug-related problems (DRP) but can also be recommendations for better prescribing and administration practices. PIs are produced daily text documents and messages forwarded to prescribers. Although they could be used retrospectively to build on safeguards for preventing DRP, the reuse of the PIs data is under-exploited.

**Objective:** The objective of this work is to train a deep learning algorithm able to automatically categorize PIs to value this large amount of data.

**Materials and Methods:** The study was conducted at the University Hospital of Strasbourg. PIs data was collected over the year 2017. Data from the first six months of 2017 was labelled by two pharmacists, who manually assigned one of the 29 possible classes from the French Society of Clinical Pharmacy classification. A deep neural network classifier was trained to learn to automatically predict the class of PIs from the processed text data.

**Results:** 27,699 labelled PIs were used to train and evaluate a classifier. The accuracy of the prediction calculated on the validation dataset was 78.0%. We predicted classes for the PIs collected in the second half of 2017. Of the 4,460 predictions checked manually, 67 required corrections. These verified data was concatenated with the original dataset to create an extended dataset to re-train the neural network. The accuracy achieved was 81.0 %, showing that the prediction process can be further improved as the amount of data increases.

**Conclusions:** PIs classification is beneficial for assessing and improving pharmaceutical care practice. Here we report a high-performance automatic classification of PIs based on deep learning that could find a place in highlighting the clinical relevance of the drug prescription review performed daily by hospital pharmacists.

## 1. INTRODUCTION

Medication review (MedRev) of drug prescriptions is a critical step to optimize prescribing, prevent drug-related harm and improve patients safety [1]. MedRev allows to intercept drug-related problems (DRP) or mitigate medication utilization [2–4]. DRP are a composite set of events (prescriptions errors, inadequate monitoring …) involving drug therapy and can refer to any events or circumstances that actually or potentially interfere with desired health outcomes [5–7]. DRP must trigger pharmaceutical interventions (PIs). PIs are defined as any proposals for treatment modification initiated by the pharmacist [8–11]. PIs are proposed during the medication order review process - and must be triggered when a DRP is detected. But PIs also correspond to recommendations for best practices in prescribing and administration [12,13]. PIs are generally well documented, in the form of text documents and messages forwarded to prescribers. But these data are rarely reused although they could contribute to retrospectively assess guidelines and safeguards for preventing drug-related problems.

To be useful for this purpose, it is necessary to classify PIs in homogeneous categories relevant and informative on DRP or on the actions proposed to prescribers. Many countries have developed their own DRP classification system [14–19]. But most of these classifications are focusing on DRP, missing the additional targets of PIs. The French Society of Clinical Pharmacy (SFPC) has adopted a 11 categories (derived in a 29 categories and sub-categories) classification for PIs [20,21]. This is currently the preferred method for recording PIs used both in daily pharmaceutical care practice and research in France.

Over the past decade, the implementation of prescription assistance software in has facilitated the deployment of MedRev as prescriptions have been integrated into massive electronic medical record data. Several recent works capitalized on machine learning algorithm processing this flux of data to facilitate the MedRev process [22–26] or their analysis [27,28]. Once proposed by pharmacists, PIs are also captured, logged and archived as part of the patient medical record. These data constitute information-rich data-marts - although poorly reused - which could provide a unique opportunity to develop expert artificial intelligence-based tools to help pharmacists to retrospectively classify PIs, analyse PIs patterns and ultimately alert them to certain risky prescribing and administration attitudes.

In this context, the objective of this study was to develop a deep neural network classifier able to predict from the French text documents of PIs the most appropriate PIs classes.

## 2. MATERIALS AND METHODS

### 2.1. Dataset

The study was conducted in a large, public, University Hospital (over 2,400 beds) in Strasbourg, France, that provides surgical, medical, research and teaching activities. Data were collected over a full calendar year period (January through December 2017) directly from the hospital information system. Data corresponded to the PIs generated on prescriptions of all adult inpatients at the exclusion of those admitted in intensive care units, obstetrics and gynaecology, or psychiatry units. The extracted data included pharmacists comments and documents describing (in French) the reason for the PIs together with messages forwarded to the prescribers.

### 2.2. Classification of pharmacist interventions

The classification of PIs was based on the externally validated SFPC classification [20]. The hierarchical structure of the SFPC classification defines 28 classes and subclasses for PIs (see table 1).

**Table 1:** French Society of Clinical Pharmacy classification of pharmacist interventions [13].

| Code | Pharmacist intervention |
| --- | --- |
| **1** | Non conformity to guidelines or contra-indication |
| **1.1** | Non conformity of the drug choice compared to the Formulary |
| **1.2** | Non conformity of the drug choice compared to the guidelines |
| **1.3** | Physio-pathologic contra-indication |
| **2** | Untreated indication |
| **2.1** | Valid indication without a drug |
| **2.2** | A drug is missing after transfer |
| **2.3** | The patient is missing a pre-medication or a prophylactic treatment |
| **2.4** | A synergic or corrective drug should be associated |
| **3** | Sub-therapeutic dosage |
| **3.1** | Dose too low for this specific patient |
| **3.2** | Length of the treatment too short |
| **4** | Overdosage |
| **4.1** | Supra-therapeutic posology |
| **4.2** | Duplicate prescription |
| **5** | Drug without indication |
| **5.1** | No justified indication for the drug |
| **5.2** | The drug is being prescribed for a too long period without any risks |
| **5.3** | Therapeutic redundancy |
| **6** | Drug interaction |
| **6.1** | Take into account |
| **6.2** | Precaution for use |
| **6.3** | A drug interferes with another drug and can lead to a non-adapted pharmacological impact |
| **6.4** | Contra-indication between 2 drugs |
| 6.5 | Interaction published but not integrated into Vidal © dictionary |
| 7 | Adverse drug reaction |
| 8 | Improper administration |
| 8.1 | Other route more effective or less costly for the same efficacy |
| 8.2 | The method for administration is not adequate |
| 8.3 | Inappropriate drug form |
| 8.4 | Incomplete formulation |
| 8.5 | Inappropriate timing of administration and/or repartition of doses |
| 9 | Failure to receive drug |
| 9.1 | Physico-chemical incompatibility between several injectable drugs |
| 9.2 | Compliance problem |
| 10 | Drug monitoring |
| 11 | Other |

### 2.3. Manual labelling of pharmacist interventions

PIs generated over the first six-months of 2017 were reviewed independently by two pharmacists who assigned each document a PI code number. In case of multiple possible class assignments, the one considered potentially as the most harmful for the patient was selected. If a discrepancy between the 2 pharmacists occurred, the PI labelling was re-evaluated in order to reach a consensus. This human review process allowed to label about 30,000 PIs with a unique PI code. As anticipated, labelled classes were largely unbalanced.

### 2.4. Data split

To account for the unbalanced number of data by class, a stratified split of the data was performed to build a training (85%) and a validation dataset (15%).

### 2.5. Text processing

PI documents were written as French plain text - often in the form of technical notes for prescribers and needed to be processed. Text processing was performed using R (version 4.1.0) [29]. Briefly, abbreviations and keywords have been replaced by their corresponding expressions. Accents, punctuations, numbers and duplicated documents were removed. The Porter’s stemming algorithm (package tm) was used to stem French words in the corpus of texts. Terms were tokenized as n-grams (n from 1 to 3) and a document term matrix was constructed using term frequency weighting. Data from the validation dataset were processed similarly at the exception of the very last step where only terms initially present in the training document term matrix were kept to build the validation document term matrix.

### 2.6. Deep neural network classifier

A deep neural network classifier was built from R using Keras library package (2.4.0) [30] and running on Tensorflow backend (2.5.0) [31]. The model was composed of three dense layers with Rectified Linear Unit (ReLU) activation functions and regularized using dropout to avoid overfitting. The 29 units’ output layer was activated with a softmax function. The model had about 3 × 10^6^ trainable parameters. A weighted categorical cross entropy loss function was used to account for unbalanced classes (weights were defined as the inverse of the class frequencies). The loss function was minimized using Adaptative Moment Estimation (Adam) optimizer. The model was trained using a batch size of 64 over about 10 epochs. The training time was fast and the algorithm converged smoothly. We used Accuracy (Acc = (TP+TN)/N), Specificity (Sp =TN/(TN+FP)) and Sensitivity (or recall) (Se = TP/(TP+FN) metrics for performance evaluation. For classes and subclasses where the number of cases was less than 5, performance indicators were not reported, as the interpretation for these data was poorly indicative.

The code is made available on an open repository at https://git.unistra.fr/jgodet/deep_pi

### 2.7. Ethics committee approval

The local Ethics committee (Comité d’éthique des Facultés de Médecine, d’Odontologie, de Pharmacie, des Écoles d’Infirmières, de Kinésithérapie, de Maïeutique et des Hôpitaux) approved this non-interventional and retrospective study (reference CE-2022-21).

## 3. RESULTS

### 3.1. Manual encoding of pharmacists’ interventions

A total of 27,699 unique text documents of PIs were extracted for the first six-months of 2017. For each document, a specific PI code was attributed independently by the two pharmacists. The amount of divergent coding between the two ratters were n= 1,006 discrepancies (3.6%) for which a consensus labels were determined. It is important to remind that a single code was attributed to every document. Three subclasses (codes 2.3, 6.5, 9.2) were not observed in the dataset. In order to make the algorithm able to learn these classes, we manually added credible synthetic comments proposed by clinical pharmacists, resulting finally in 27,721 unique documents (table 2).

**Table 2:** Performance of the initial algorithm in terms of average predictive score, sensitivity, and specificity for the different classes and subclasses of PIs.

| Code | Pharmacist intervention | Initial classifier<br>(Global Accuracy: **78.0%**) |  |  |
| --- | --- | --- | --- | --- |
|  |  | Initial dataset<br>n | Validation dataset<br>n<br>(% of the PIs) | Prediction Performance |
| **1** | **Non conformity to guidelines or contra-indication** |  |  |  |
| **1.1** | Non conformity of the drug choice compared to the Formulary | 11,935 | 1,790<br>(43.2%) | Avg pred score : 0.96<br>Se : 0.93<br>Sp : 0.94 |
| **1.2** | Non conformity of the drug choice compared to the guidelines | 1,434 | 215<br>(5.0%) | Avg pred score : 0.71<br>Se : 0.48<br>Sp : 0.97 |
| **1.3** | Physio-pathologic contra-indication | 166 | 24<br>(0.6%) | Avg pred score : 0.69<br>Se : 0.37<br>Sp : 0.99 |
| **2** | **Untreated indication** |  |  |  |
| **2.1** | Valid indication without a drug | 90 | 13<br>(0.3%) | Avg pred score : 0.66<br>Se : 0.31<br>Sp : 0.99 |
| **2.2** | A drug is missing after transfer | 328 | 49<br>(1.2%) | Avg pred score : 0.92<br>Se : 0.96<br>Sp : 0.99 |
| **2.3** | The patient is missing a pre-medication or a prophylactic treatment | 0*(12) | 1<br>(0.0%) | Avg pred score : 0.63<br>Se : NA<br>Sp : NA |
| **2.4** | A synergic or corrective drug should be associated | 467 | 70<br>(1.7%) | Avg pred score : 0.88<br>Se : 0.78<br>Sp : 0.99 |
| **3** | **Subtherapeutic dosage** |  |  |  |
| **3.1** | Dose too low for this specific patient | 1,108 | 166<br>(4.0%) | Avg pred score : 0.72<br>Se : 0.31<br>Sp : 0.98 |
| **3.2** | Length of the treatment too short | 29 | 4<br>(0.1%) | Avg pred score : 0.41<br>Se : NA<br>Sp : NA |
| **4** | **Overdosage** |  |  |  |
| **4.1** | Supra-therapeutic posology | 2,824 | 423<br>(10.2%) | Avg pred score : 0.85<br>Se : 0.74<br>Sp : 0.96 |
| **4.2** | Duplicate prescription | 787 | 118<br>(2.8%) | Avg pred score : 0.86<br>Se : 0.81<br>Sp : 0.99 |
| **5** | **Drug without indication** |  |  |  |
| **5.1** | No justified indication for the drug | 585 | 87<br>(2.1%) | Avg pred score : 0.79<br>Se : 0.67<br>Sp : 0.99 |
| **5.2** | The drug is being prescribed for a too long period without any risks | 39 | 5<br>(0.1%) | Avg pred score : 0.44<br>Se : 0.80<br>Sp : 0.99 |
| **5.3** | Therapeutic redundancy | 340 | 51<br>(1.2%) | Avg pred score : 0.75<br>Se : 0.43<br>Sp : 0.99 |
| **6** | **Drug interaction** |  |  |  |
| **6.1** | Take into account | 4 | 0<br>(0.0%) | Avg pred score : NA<br>Se : NA |
|  |  |  |  | Sp NA |
| **6.2** | Precaution for use | 409 | 61<br>(1.5%) | Avg pred score : 0.84<br>Se : 0.79<br>Sp : 0.99 |
| **6.3** | A drug interferes with another drug and can lead to a non-adapted pharmacological impact | 108 | 16<br>(0.4%) | Avg pred score : 0.76<br>Se : 0.75<br>Sp : 0.99 |
| **6.4** | Contra-indication between 2 drugs | 647 | 97<br>(2.3%) | Avg pred score : 0.87<br>Se : 0.81<br>Sp : 0.99 |
| **6.5** | Interaction published but not integrated into Vidal © dictionary | 0*(5) | 0<br>(0.0%) | Avg pred score : NA<br>Se : NA<br>Sp : NA |
| **7** | Adverse drug reaction | 12 | 1<br>(0.0%) | Avg pred score : 0.34<br>Se : NA<br>Sp : NA |
| **8** | Improper administration |  |  |  |
| **8.1** | Other route more effective or less costly for the same efficacy | 68 | 10<br>(0.2%) | Avg pred score : 0.92<br>Se : 0.90<br>Sp : 1.0 |
| **8.2** | The method for administration is not adequate | 449 | 67<br>(1.6%) | Avg pred score : 0.86<br>Se : 0.78<br>Sp : 0.99 |
| **8.3** | Inappropriate drug form | 1,246 | 186<br>(4.5%) | Avg pred score : 0.86<br>Se : 0.77<br>Sp : 0.99 |
| **8.4** | Incomplete formulation | 460 | 69<br>(1.7%) | Avg pred score : 0.86<br>Se : 0.77<br>Sp : 0.99 |
| **8.5** | Inappropriate timing of administration and/or repartition of doses | 1,298 | 194<br>(4.7%) | Avg pred score : 0.85<br>Se : 0.74<br>Sp : 0.98 |
| **9** | Failure to receive drug |  |  |  |
| **9.1** | Physico-chemical incompatibility between several injectable drugs | 26 | 3<br>(0.1%) | Avg pred score : 0.51<br>Se : NA<br>Sp : NA |
| **9.2** | Compliance problem | 0*(5) | 0<br>(0.0%) | Avg pred score : NA<br>Se : NA<br>Sp : NA |
| **10** | Drug monitoring | 875 | 131<br>(3.2%) | Avg pred score : 0.85<br>Se : 0.66<br>Sp : 0.99 |
| **11** | Other | 1,965 | 294<br>(7.1%) | Avg pred score : 0.77<br>Se : 0.59<br>Sp : 0.97 |

Avg pred score: Average predictive score probability (mean of the highest softmax probabilities);
Se: Sensitivity; Sp: Specificity
0(n)\*: PIs not observed in the dataset (n= number of credible synthetic comments added)

### 3.2. Pharmacist interventions processing

The 27,721 PI documents were split (stratification on classes and subclasses) to form a 23,576 (85%) training dataset and a 4,145 (15%) validation dataset. In the training set, processed comments (stemmed and cleaned) were typically short with a median number of 82 characters (IQR = [59-115], min = 3, max = 1,153), corresponding to a median number of 13 words or expressions. Comments were tokenized using n-grams (n from 1 to 3). We used the full set of documents in the training set to generate a (178,680 × 23,576) term-document matrix based on term frequency weighting. The size of this initial matrix was reduced to (6,356 × 23,576) after removing sparse terms. This (6,356 × 23,576) term-document matrix was transposed in a (23,576 × 6,356) training dataset. Using a similar pre-processing, we obtained a (4,145 × 6,356) validation dataset.

### 3.3. Deep neural network classifier

The training dataset and its corresponding 23,576 ground truth PI classes and subclasses were fed to a deep neural network composed of 3 dense layers including a 29-classes and subclasses output layer activated by a softmax function. The loss function was weighted by the inverse of the class frequencies of the training labels to account for unbalanced class representations in the training dataset. The class prediction accuracy calculated on the unseen validation dataset was 78.0%. A tabulated description of data by PIs classes and subclasses is presented in Table 2 (initial algorithm). Classes and subclasses sensibilities and specificities ranged from 0.31 to 0.96 and from 0.94 to 1.0, respectively. The output of the softmax function, normalizing the output of the network last layer to a probability distribution, was used to calculate a predictive probability score for each class label. The predicted probability scores were largely asymmetric, (mean = 0.879, median = 0.999) with 2,626 out of the 4,145 predicted scores (63.3%) being larger than 0.95. As expected, the global accuracy associated to predictions with scores larger than 0.95 was largely increased as compared to the overall prediction, reaching 94.7%.

In order to further demonstrate the classification capacity of this algorithm, we predicted the classes and subclasses for all the pharmacists’ comments collected between July and December 2017. A term-document matrix was generated and fed to the trained neural network to retrieve class predictions. All class predictions with a probability prediction score larger or equal to 0.95 and a code differing from the subcategory “non-conformity of the drug choice compared to guidelines – code 1.1” (this subclass can be easily automatically checked using drugs dictionaries) were selected and manually checked by a pharmacist. Amongst the 4,460 predictions checked, 67 needed to be corrected, corresponding to an error rate of about 1.5%. Finally, this dataset was concatenated with the original one to create an extended dataset (15% larger than the original) that we split as previously into a training and a validation dataset. The network was trained again from scratch on this expanded dataset. The resulting global accuracy reached then 81.0% (+3.0 percent points) (see table 3 - improved algorithm).

**Table 3:** Performance of the algorithm with extended training set in terms of average predictive score, sensitivity, and specificity for the different classes and subclasses of PIs.

| Code | Pharmacist intervention | Classifier with extended training dataset<br>(Accuracy: 81.0%) |  |  |
| --- | --- | --- | --- | --- |
|  |  | Extended dataset<br>(n) | Validation dataset<br>(n)<br>(% of the PIs) | Prediction Performance |
| **1** | Non conformity to guidelines or contra-indication |  |  |  |
| **1.1** | Non conformity of the drug choice compared to the Formulary | 11935 | 2.388<br>(37.2%) | Avg pred score : 0.95<br>Se : 0.93<br>Sp : 0.96 |
| **1.2** | Non conformity of the drug choice compared to the guidelines | 1515 | 302<br>(4.7%) | Avg pred score : 0.70<br>Se : 0.52<br>Sp : 0.97 |
| **1.3** | Physio-pathologic contra-indication | 166 | 33<br>(0.5%) | Avg pred score : 0.62<br>Se : 0.42<br>Sp : 0.99 |
| **2** | Untreated indication |  |  |  |
| **2.1** | Valid indication without a drug | 90 | 18<br>(0.3%) | Avg pred score : 0.66<br>Se : NA<br>Sp : 0.99 |
| **2.2** | A drug is missing after transfer | 563 | 112<br>(1.7%) | Avg pred score : 0.95<br>Se : 0.95<br>Sp : 0.99 |
| **2.3** | The patient is missing a pre-medication or a prophylactic treatment | 12 | 2<br>(0.0%) | Avg pred score : 0.68<br>Se : NA<br>Sp : NA |
| **2.4** | A synergic or corrective drug should be associated | 599 | 119<br>(1.9%) | Avg pred score : 0.89<br>Se : 0.82<br>Sp : 0.99 |
| **3** | Subtherapeutic dosage |  |  |  |
| **3.1** | Dose too low for this specific patient | 1208 | 242<br>(3.8%) | Avg pred score : 0.72<br>Se : 0.52<br>Sp : 0.98 |
| **3.2** | Length of the treatment too short | 29 | 5<br>(0.1%) | Avg pred score : 0.56<br>Se : NA<br>Sp : NA |
| **4** | Overdosage |  |  |  |
| **4.1** | Supra-therapeutic posology | 3849 | 767<br>(11.9%) | Avg pred score : 0.84<br>Se : 0.72<br>Sp : 0.98 |
| **4.2** | Duplicate prescription | 1399 | 277<br>(4.3%) | Avg pred score : 0.90<br>Se : 0.89<br>Sp : 0.99 |
| **5** | Drug without indication |  |  |  |
| **5.1** | No justified indication for the drug | 679 | 136<br>(2.1%) | Avg pred score : 0.73<br>Se : 0.65<br>Sp : 0.99 |
| **5.2** | The drug is being prescribed for a too long period without any risks | 39 | 7<br>(0.1%) | Avg pred score : 0.46<br>Se : 0.86<br>Sp : 1.0 |
| **5.3** | Therapeutic redundancy | 343 | 70 | Avg pred score : 0.73 |
|  |  |  | (1.1%) | Se : 0.44<br>Sp : 0.99 |
| **6** | **Drug interaction** |  |  |  |
| **6.1** | Take into account | 4 | 0<br>(0.0%) | Avg pred score : NA<br>Se : NA<br>Sp : NA |
| **6.2** | Precaution for use | 738 | 147<br>(2.3%) | Avg pred score : 0.91<br>Se : 0.87<br>Sp : 0.99 |
| **6.3** | A drug interferes with another drug and can lead to a non-adapted pharmacological impact | 108 | 21<br>(0.3%) | Avg pred score : 0.70<br>Se : 0.48<br>Sp : 0.99 |
| **6.4** | Contra-indication between 2 drugs | 865 | 173<br>(2.7) | Avg pred score : 0.95<br>Se : 0.92<br>Sp : 0.99 |
| **6.5** | Interaction published but not integrated into Vidal © dictionary | 5 | 1<br>(0.0%) | Avg pred score : 0.98<br>Se : NA<br>Sp : NA |
| **7** | **Adverse drug reaction** | 12 | 2<br>(0.0%) | Avg pred score : 0.40<br>Se : NA<br>Sp : NA |
| **8** | **Improper administration** |  |  |  |
| **8.1** | Other route more effective or less costly for the same efficacy | 132 | 26<br>(0.4%) | Avg pred score : 0.95<br>Se : 0.88<br>Sp : 0.99 |
| **8.2** | The method for administration is not adequate | 589 | 117<br>(1.8%) | Avg pred score : 0.86<br>Se : 0.87<br>Sp : 0.99 |
| **8.3** | Inappropriate drug form | 1527 | 304<br>(4.7%) | Avg pred score : 0.87<br>Se : 0.81<br>Sp : 0.99 |
| **8.4** | Incomplete formulation | 596 | 119<br>(1.9%) | Avg pred score : 0.87<br>Se : 0.84<br>Sp : 0.99 |
| **8.5** | Inappropriate timing of administration and/or repartition of doses | 1821 | 364<br>(5.7%) | Avg pred score : 0.89<br>Se : 0.83<br>Sp : 0.98 |
| **9** | **Failure to receive drug** |  |  |  |
| **9.1** | Physico-chemical incompatibility between several injectable drugs | 26 | 5<br>(0.1%) | Avg pred score : 0.53<br>Se : NA<br>Sp : NA |
| **9.2** | Compliance problem | 5 | 1<br>(0.0%) | Avg pred score : 0.89<br>Se : NA<br>Sp : NA |
| **10** | **Drug monitoring** | 1158 | 231<br>(3.6%) | Avg pred score : 0.85<br>Se : 0.77<br>Sp : 0.99 |
| **11** | **Other** | 2169 | 436<br>(6.8%) | Avg pred score : 0.76<br>Se : 0.66<br>Sp : 0.98 |

Avg pred score: Average predictive score probability (mean of the highest softmax probabilities); Se: Sensitivity; Sp: Specificity

## 4. DISCUSSION

Here we study present a simple pipeline including a robust deep neural network classifier to categorize pharmacist interventions from the raw text data used to document them. To our knowledge, no other deep learning PI classification system has been described in the literature to date. The classification relied on the external validated SFPC classification - currently the most widely used classification method of PIs in France [20]. It contains explicit definitions for each category of PIs to reduce ambiguity in coding a problem. The 29 classes and subclasses of the initial dataset were unbalanced, with an over- representation of the situations corresponding to “non-conformity of the drug choice compared to the formulary” and “non-conformity of the drug choice compared to the guidelines”. On the opposite, there was an under-representation of classes such as “adverse drug reaction” (∼4/10,000) or “failure to receive drug” (∼1/1,000) - in full line with the fact that these events are rare. But as compared to Bedouch et al. [32], where “adverse drug reaction” or “failure to receive a drug” accounted for 4.3% and 0.8% of their 34,522 interventions, respectively, we observed clearly less events in these two categories - possibly because they were focusing on DRP. Indirectly it shows that PIs are not strictly limited to DRP issues in the prescription review process.

The SFPC classification seems well adapted to describe pharmacist interventions in our hospital. Indeed, the class named “other” represented only 7.1% of all PIs of the initial dataset. This likely includes possible coding difficulties for the experts due to a lack of details in the documentation or because of complexity of interpretations. This has to be compared with some studies in which the “other” class represented up to 39% of all DRP [15].

Regarding performance, a global accuracy of 78% was achieved on the validation dataset. The model performed worst for subclasses such as, “physio-pathologic contra-indication”, “valid indication without drug”, “dose too low for this specific patient”. Several explanations can be given - including insufficient number of cases in the initial dataset or more linguistically complex documents that may require some inference to understand which class the entities involved are related to.

Algorithmic prediction errors in the external validation dataset (second six-months of 2017) spotted by an expert pharmacist were low (1.5% of the predictions with a probability larger or equal to 0.95 and a code differing from the subcategory “non-conformity of the drug choice compared to guidelines – code 1.1”). A detailed manual analysis of these errors showed that they corresponded to complex clinical situations where multiple PIs could be encoded.

Incorporating additional validated data from the second half of 2017 (see figure 1) into an expanded dataset improved the performance of the classifier (accuracy reached 81%). This clearly shows that the prediction process has the potential to improve further with an increase in the amount of data and strengthens the validity of the results.

**Figure 1:**
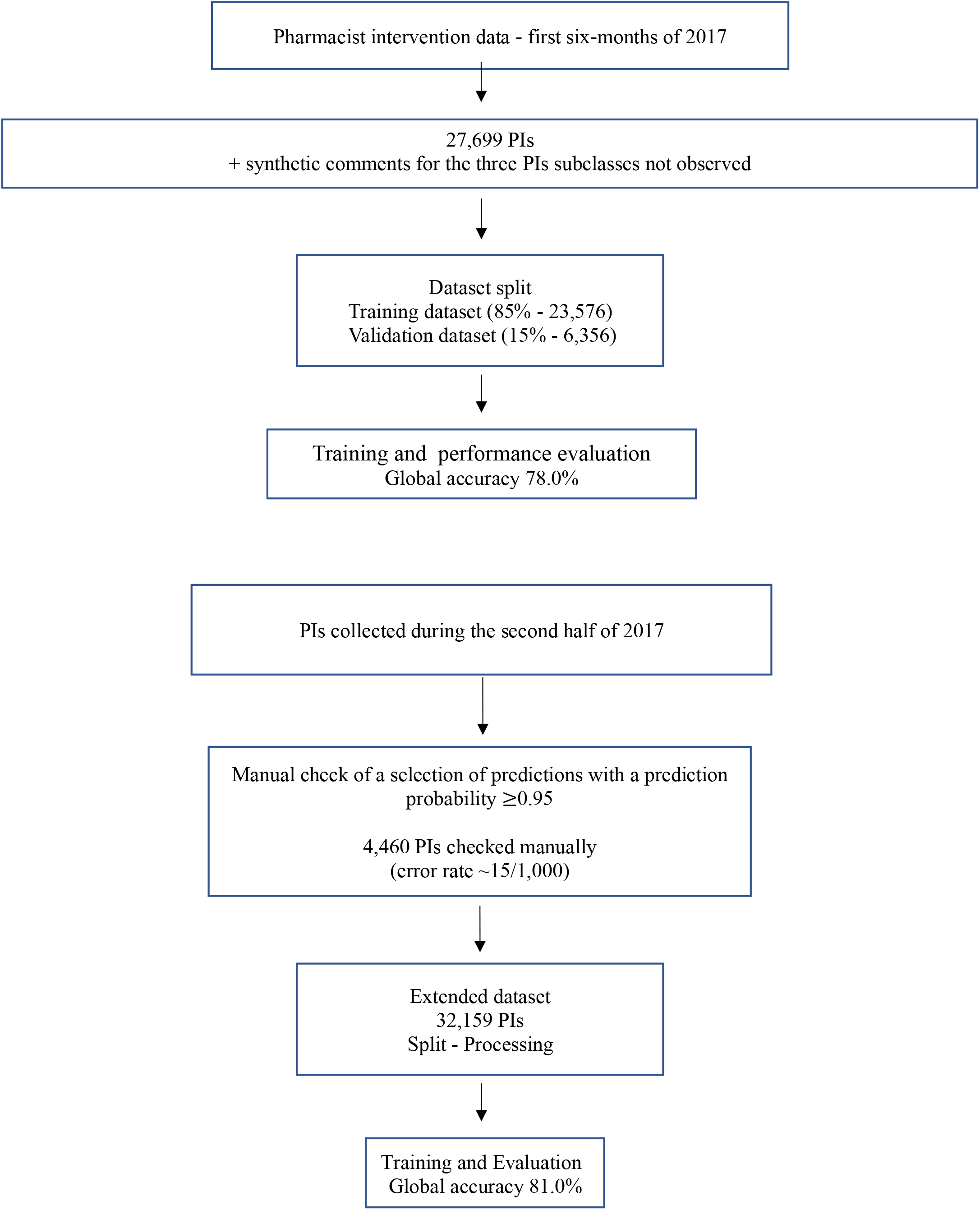
Flowchart summarizing the different steps that led to the training and evaluation of the classifier

This study has several limitations. Our findings are currently limited in scope as the study was conducted in a single French hospital setting. Its generalization properties to other institutions has not been explored. Adult patients in intensive care units, obstetrics and gynaecology, and psychiatry together with children patients were not included. Consequently, evidence of the accuracy of the algorithm to classify PIs in these targeted populations has yet to be demonstrated.

The future objectives will be to exploit this deep neural network algorithm to perform large descriptive analysis of PIs and DRP intercepted through MedRev with the aim to value the work of clinical pharmacists. The description of PI classes, automatically generated by the algorithm, will be crucial to increase knowledge on the types and frequencies of PI or DRP observed in different clinical settings. Particular attention will be paid to the prevalence rates of PI by drug classes, by patient profiles (age, number of drugs prescribed) or medical specialities. Although this type of analysis will focus on the drug related problems spotted by pharmacists but not on their cause or consequence, it will be nevertheless useful to alert medical and pharmaceutical teams about certain prescribing patterns and draw the attention on practices that could be improved. As PIs can be influenced by the pharmacists’ experience, it would be interesting in a next work to compare the PIs pattern issued from junior as compared to those of experienced senior pharmacists. The classifier could be used to check differences between junior and senior pharmacist practice and to assess the hospital’s pharmacy quality policy requiring that all residents and newly hired pharmacists undergo specific training to be authorized to perform PI and write comments.

## CONCLUSION

PIs classification is beneficial for assessing and improving pharmaceutical care practice. It should be systematized as part of the care process. Here we report a high performance automatic PIs classification based on deep learning that could find a crucial place for highlighting the clinical relevance of drug prescription reviews performed on a daily basis by hospital pharmacists

## Data Availability

All data produced in the present study are available upon reasonable request to the authors

## AUTHOR CONTRIBUTIONS

Conceptualization: AA, JG, EJ, BG and BM;

Data labelling : AA, EJ

Data acquisition and curation: AA, JG;

Formal analysis: AA, JG;

Software coding: JG;

Supervision: BG, BM;

Visualization: AA;

Writing of original draft: AA, JG, BM;

Writing review and editing: AA, JG, EJ, BG and BM

All authors approved the final version of the work to be published; and agreed to be accountable for all aspects of the work in ensuring that questions related to the accuracy or integrity of any part of the work are appropriately investigated and resolved.

## ACKNOWLEDGEMENTS

The authors thank Dr. T. Fabacher and Dr. V. Wenger for their valuable assistance in querying the data in the hospital information system.

## SUMMARY TABLE

**what was already known on the topic**

- Pharmacist Interventions classification is beneficial for assessing and improving pharmaceutical care practice
- Although PIs could be used to build on retrospectively safeguards for preventing DRP, PIs data are underexploited.

**what this study added to our knowledge**

- Here we report a high performance automatic PIs classification based on deep learning
- Analysis of reliable PIs class may find a crucial place for highlighting the clinical relevance of drug prescription reviews performed on a daily basis by hospital pharmacists

